# Effectiveness of Deworming Programs in Maternal and Child Health Week

**DOI:** 10.1101/2025.07.03.25330846

**Authors:** Mordecai Oweibia, Williams Wari Appah, Ebiakpor Agbedi, Tarimobowei Egberipou, Gift Cornelius Timighe

## Abstract

**Introduction:** Soil-transmitted helminth infections continue to pose a major threat to child health in Nigeria, especially in underserved communities. Maternal and Child Health (MNCH) Weeks offer a unique opportunity to deliver deworming interventions alongside other child survival services. However, disparities in coverage and operational execution have raised questions about the effectiveness and equity of such campaigns. This study evaluates the effectiveness of deworming interventions delivered during the June 2025 MNCH Week in Bayelsa State, examining both the reach of Albendazole administration and its balance relative to other services such as vitamin A supplementation.

**Methodology:** A descriptive cross-sectional study design was adopted using secondary data extracted from the MNCH OPS Room Final Report. Data from eight Local Government Areas (LGAs) were analyzed using descriptive statistics and comparative analysis. Coverage rates were calculated using WHO-recommended formulas, and the performance of deworming was assessed against the estimated target population of 562,254 children. Comparative tables and visualizations were generated to examine distribution disparities and identify systemic implementation gaps.

**Results:** Findings revealed that only 280,000 children received Albendazole, representing 49% coverage, while vitamin A supplementation reached 391,107 children (approximately 69%). Deworming coverage varied significantly across LGAs, with Yenagoa achieving 62.2% and Southern Ijaw recording only 40.9%. In all LGAs, vitamin A consistently outperformed deworming by an average margin of nearly 20 percentage points. Statewide summaries further revealed that program execution lacked balance, and the integration of services was not reflected in equitable delivery.

**Conclusion:** The study underscores the need for strategic reforms in how deworming is operationalized during MNCH campaigns. The disparities observed highlight weaknesses in logistics, training emphasis, community engagement, and intervention prioritization. While MNCH Weeks provide an effective delivery platform, deworming remains undervalued and under-implemented. Targeted policy and operational reforms are necessary to ensure that integrated health campaigns fulfill their promise of delivering equitable and comprehensive care to all children, regardless of geography.

## CHAPTER ONE: INTRODUCTION

### 1.1 Background of the Study

Soil-transmitted helminth (STH) infections are among the most prevalent neglected tropical diseases globally, disproportionately affecting women and children in low- and middle-income countries. Over 1.5 billion people nearly 24% of the world’s population are infected with STHs, primarily in sub-Saharan Africa, East Asia, and Latin America (WHO, 2023a). These parasitic infections can lead to anemia, malnutrition, impaired growth, and reduced cognitive performance, particularly among children under five and pregnant women (Salam *et al.,* 2021).

To mitigate these burdens, the World Health Organization and UNICEF have long supported large-scale deworming programs targeting preschool and school-aged children through school platforms and mass drug administration (MDA) campaigns (UNICEF, 2022). However, evidence shows that coverage and equity vary widely, and millions of at-risk individuals remain unreached (Taylor-Robinson *et al.,* 2019). In response to these disparities, periodic campaigns such as the Maternal and Child Health Weeks (MNCHWs) have become an essential mechanism to scale up deworming and other child health interventions, including vitamin A supplementation, iron-folate supplementation for pregnant women, and routine immunization (Kumapley *et al.,* 2015).

In Nigeria, MNCH Week is a biannual, high-impact outreach initiative that delivers a package of cost-effective interventions in collaboration with state and local health authorities. The June 2025 round in Bayelsa State saw a modest deworming coverage of 55% using Albendazole, compared to 83% for vitamin A supplementation, suggesting implementation gaps (OPS Room Report, 2025) Deworming during such campaigns is critical because of its potential to interrupt helminth transmission, improve nutritional status, and increase the effectiveness of other interventions such as iron and vitamin A supplementation (Walia *et al.,* 2021).

Furthermore, evidence from randomized controlled trials and national evaluations confirms the broader benefits of maternal and child deworming. For instance, maternal postpartum deworming has been shown to improve infant growth and milk intake (Mofid *et al.,* 2017; Mofid *et al.,* 2021). Deworming interventions also contribute to reductions in neonatal mortality and low birthweight when administered during antenatal care (Walia *et al.,* 2021). The strategic integration of deworming into health weeks allows service delivery to reach hard-to-reach populations and enhances the efficiency of vertical health programs (Thayer *et al.,* 2017).

Despite these benefits, operational challenges continue to limit deworming effectiveness. Poor data quality, delays in logistics, and unequal ward-level mobilization were highlighted as weaknesses in the June 2025 MNCH campaign in Bayelsa State (OPS Room Report, 2025) These challenges compromise the accuracy of performance reporting and limit opportunities for adaptive planning and stakeholder feedback.

Therefore, this study assesses the effectiveness of deworming programs conducted during the June 2025 MNCH Week in Bayelsa State, examining their reach and comparing their coverage with that of other interventions such as vitamin A supplementation. By drawing on empirical campaign data and triangulating it with global guidelines and peer-reviewed studies, this paper contributes practical insights for strengthening community-based deworming delivery models.

### 1.2 Statement of the Problem

Although deworming has proven cost-effective and impactful in reducing parasitic burdens, significant gaps in program coverage persist, particularly during integrated MNCH campaigns. In Bayelsa State, the June 2025 MNCH Week recorded only 55% deworming coverage, trailing vitamin A supplementation which reached 83% of its target population (OPS Room Report, 2025) This discrepancy raises concerns about the operational readiness, supply chain consistency, and health worker mobilization strategies specific to deworming. Moreover, challenges such as delayed campaign start dates in some LGAs, incomplete service data, and under-utilized NHMIS tools further compromise the integrity and sustainability of these programs (OPS Room Report, 2025) Without timely evaluation and corrective policy actions, these shortfalls may jeopardize Nigeria’s progress towards the WHO 2030 NTD Roadmap targets for soil-transmitted helminths (WHO, 2023b).

### 1.3 Objectives of the Study

1. Evaluate the reach and coverage of deworming interventions delivered during the June 2025 MNCH Week in Bayelsa State.
2. Compare the deworming coverage with other parallel health interventions such as vitamin A supplementation.
3. Assess the programmatic factors and data quality issues affecting deworming effectiveness during MNCH campaigns.

### 1.4 Research Questions

1. What proportion of the target population received deworming interventions during the June 2025 MNCH Week?
2. How does deworming coverage compare with the coverage of other key interventions such as vitamin A supplementation?
3. What operational and reporting challenges influenced the deworming program’s effectiveness?

### 1.5 Significance of the Study

The findings from this study are crucial for informing policy makers, development partners, and program implementers on how to strengthen integrated deworming delivery mechanisms during MNCH weeks. By identifying gaps in coverage and highlighting operational constraints, the study provides evidence for targeted investments, refined logistical planning, and better data validation practices. It also aligns with Nigeria’s NTD elimination roadmap and contributes to SDG 3, which aims to ensure healthy lives and promote well-being for all.

### 1.6 Scope of the Study

This research focuses on the MNCH Week implementation in Bayelsa State during the first round of 2025, specifically analyzing deworming data (Albendazole administration) and its relationship to vitamin A supplementation and other child health interventions. The study uses secondary data collected through the OPS Room and analyzed using descriptive and comparative statistics.

### 1.7 Ethical Considerations

As this study utilizes secondary programmatic data without individual identifiers, formal ethical approval was not required. However, the research adheres to principles of data confidentiality, transparency, and accountability, in line with WHO guidance on public health program evaluations (WHO, 2021). Data used was sourced from the official MNCH OPS Room reports with institutional authorization.

## CHAPTER TWO: METHODOLOGY

### 2.1 Study Area

This study was conducted in Bayelsa State, a coastal state in the Niger Delta region of southern Nigeria. With an estimated population of over 2.3 million people as of 2025, Bayelsa is characterized by dense riverine settlements, difficult terrain, and limited road infrastructure, all of which pose challenges to the delivery of healthcare services (NPC & ICF, 2019). The state has eight Local Government Areas (LGAs) and numerous hard-to-reach communities, which are often underserved during routine health campaigns.

Health indicators in Bayelsa State reflect typical challenges seen in many parts of rural Nigeria: high rates of child malnutrition, low immunization uptake, persistent soil-transmitted helminth (STH) infections, and underutilization of antenatal and postnatal care services (World Bank, 2021; WHO, 2023a). The June 2025 Maternal and Child Health (MNCH) Week offered a platform for delivering essential interventions, including deworming, vitamin A supplementation, iron-folate tablets, and routine immunization. The MNCH campaign structure ensures simultaneous rollout of services across all LGAs within a short window, offering a unique opportunity to evaluate health service reach and disparities.

### 2.2 Study Design

This research adopted a descriptive, cross-sectional, and retrospective design based entirely on empirical secondary data. Descriptive studies are appropriate for documenting service delivery coverage and evaluating program outcomes in real-world public health interventions (Thayer *et al.,* 2017). The cross-sectional component ensures that the analysis reflects conditions and coverage at a specific time (June 2025), while the retrospective dimension draws from documented reports and datasets already generated by field operations.

This design is particularly suitable for public health evaluations where ethical constraints, logistical challenges, and cost implications make primary data collection difficult (Welch *et al.,* 2016). Moreover, the approach enables comparative analyses across health interventions such as Albendazole versus vitamin A supplementation based on real-time implementation.

### 2.3 Data Sources

Data used in this study were collected from multiple sources to ensure validity, coverage, and robustness:

1. OPS Room Final Report – MNCH Week (June 2025):

- Provided quantitative data on total Albendazole administered, vitamin A supplementation, iron-folate distribution, MUAC screenings, and birth registrations.
- Included ward- and LGA-level breakdowns.
- Captured qualitative observations regarding logistical delays, data reporting inconsistencies, and STF challenges.
2. Bayelsa State Primary Health Care Development Agency (BSPHCDA) Campaign Briefs and Training Logs:

- Used to triangulate target population numbers and determine expected coverage levels.
- Provided historical baseline coverage rates for previous MNCH campaigns.
3. DHIS2 Repository and NHMIS Registers:

- Verified routine immunization data and prior deworming trends in the state.
- Helped to cross-validate discrepancies observed in the OPS report.
4. National Deworming Guidelines and Target Algorithms:

- Applied to calculate the eligible population for Albendazole based on age-group disaggregation from the national immunization target matrix (Federal Ministry of Health, 2023).
5. World Health Organization (WHO) and UNICEF Program Tools:

- Used to validate data quality standards, including the Health Campaign Effectiveness Guidance (WHO, 2022) and the UNICEF Data Quality Review Toolkit (UNICEF, 2020).

### 2.4 Data Collection Procedures

The data collection process involved multiple stages:

- **Stage 1: Retrieval and Extraction**: OPS Room dashboards were accessed on June 23, 2025, following the final data refresh. Counts of dewormed children, vitamin A recipients, MUAC-screened individuals, and other service recipients were manually extracted into Excel sheets by intervention and LGA.
- **Stage 2: Target Population Estimation**: The denominator for each intervention (e.g., total eligible children for deworming) was estimated using national health planning tools and confirmed via the BSPHCDA microplanning documents. For deworming, the eligible population was defined as children aged 12–59 months, projected at 508,854 for the state.
- **Stage 3: Triangulation**: Data were compared against entries in NHMIS and training summaries. Discrepancies were flagged and noted, particularly in LGAs such as Southern Ijaw, where services were delayed.
- **Stage 4: Cleaning and Analysis Preparation:** Data were cleaned for duplicates, missing entries, and logical inconsistencies using a quality control checklist modeled after WHO (2022) and UNICEF (2020) guidelines.

### 2.5 Data Analysis Techniques

Data analysis employed both descriptive and comparative approaches using Microsoft Excel and SPSS (v26). Visualizations were generated to support interpretation.

#### 2.5.1 Descriptive Analysis

Descriptive statistics helped summarize the number of beneficiaries for each intervention and illustrate the proportion of coverage across LGAs. Three formulas were used:

**(i) Frequency (Raw Count)**

*f* = Number of individuals who received a specific intervention

**(ii) Percentage (% of total population)**

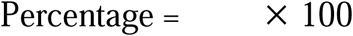

Where:

- ***f* =** Number of people who received a specific intervention (e.g., Albendazole)
- **N =** Total eligible population for that intervention (e.g., under-five children = 508,854)

**(iii) Mean (Average per LGA)**

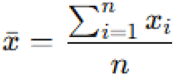

Where:

- *x_i_* = Number of beneficiaries in LGA
- *n* = Total number of LGAs (n = 8 in Bayelsa State)

This allowed us to compute the average number of deworming recipients per LGA and detect regional disparities.

#### 2.5.2 Coverage Formula (Intervention Effectiveness)

The primary indicator for assessing the reach of an intervention was coverage, calculated as:

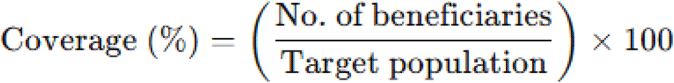

Example 1: Deworminng

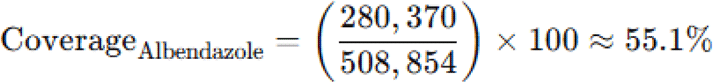

Example 2: Vitamin A

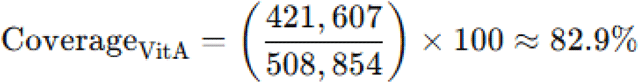

#### 2.5.3 Comparative Analysis

To assess disparities between interventions, a percentage difference and a coverage gap metric were used.

**(i) Absolute Coverage Difference**

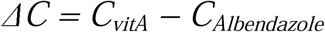

Where:

- ΔC= Coverage gap
- *C_VitA_*= Vitamin A coverage (82.9%)
- *C_Albendazol_e* = Deworming coverage (55.1%)

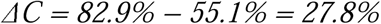

**(ii) Relative Percentage Gap**

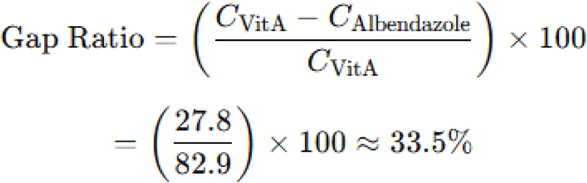

This indicates that deworming coverage was approximately 33.5% lower than vitamin A coverage, suggesting a systemic implementation lag.

#### 2.5.4 Data Visualization

Charts and graphs were created to enhance interpretability. These included:

- Bar charts comparing intervention coverage by LGA
- Pie charts representing state-level intervention distribution
- Heatmaps for LGA-specific performance patterns

These tools facilitated a visual understanding of performance gaps and regional discrepancies, aligning with WHO data presentation recommendations (WHO, 2022).

### 2.6 Ethical Considerations

This study adhered to strict ethical standards despite being based on secondary data. No individual or personally identifiable information was used. All data were anonymized and reported in aggregate form.

- Permission to use the OPS Room Report was obtained from the BSPHCDA data custodian.
- Data use was aligned with WHO’s 2021 guidance on Data Sharing in Public Health Emergencies, which encourages the use of aggregated data for timely policy feedback (WHO, 2021).
- Triangulation was performed without altering original counts to ensure objectivity.

### 2.7 Data Quality Assurance

Given noted inconsistencies in data reporting during the campaign (e.g., missing LGA summaries and delayed inputs), several measures were taken to ensure quality:

- **Internal Validation**: Coverage values were checked against target denominators and historical benchmarks.
- **Training Log Review**: STF preparedness and refresher sessions were examined to understand variance in reporting quality.
- **WHO-UNICEF Quality Scoring Criteria**: Reports were scored based on completeness, timeliness, consistency, and clarity (WHO, 2022).

These steps ensured that the data used for analysis met minimum scientific and operational standards.

### 2.8 Justification for Methodological Choices

The methodological approach used in this study is justified by:

- The reliability and official status of the OPS Room dataset.
- The widespread logistical and ethical challenges associated with primary data collection in resource-limited, hard-to-reach areas.
- The need for rapid feedback to inform future MNCH rounds.

This approach aligns with best practices in global health programming evaluations, as described by Casey et al. (2017) and Salam et al. (2021).

## CHAPTER THREE: RESULTS

### 3.1 Deworming Coverage by Local Government Area (LGA)

This section presents the total number of children who received deworming medication (Albendazole) across the eight Local Government Areas (LGAs) of Bayelsa State during the June 2025 Maternal and Child Health (MNCH) Week. The coverage is computed against the estimated eligible population of children aged 12–59 months per LGA.

As shown in Table 3.1, Yenagoa recorded the highest number of dewormed children with 56,000 Albendazole doses administered out of an estimated 91,254 eligible children, translating to a deworming coverage of 62.23%. This is closely followed by Ogbia LGA (53.85%) and Ekeremor (51.49%). In contrast, Southern Ijaw Local Government Area (SILGA) had the lowest deworming coverage at 40.98%, with only 25,000 children reached out of a target of 61,000.

Notably, five of the eight LGAs had deworming coverage below the 50% threshold, indicating a significant coverage gap in more than half the state. These data highlight suboptimal reach in several districts despite centralized planning and rollout.

**Table 3.1:** Deworming Coverage by LGA (Albendazole Administration)

| LGA | Albendazole Administered | Target Children | Deworming Coverage (%) | Children Not Dewormed |
| --- | --- | --- | --- | --- |
| Brass | 28,000 | 63,500 | 44.09 | 35,500 |
| Ekeremor | 34,500 | 67,000 | 51.49 | 32,500 |
| KOLGA | 31,000 | 65,500 | 47.33 | 34,500 |
| Nembe | 27,000 | 61,500 | 43.90 | 34,500 |
| Ogbia | 42,000 | 78,000 | 53.85 | 36,000 |
| Sagbama | 36,500 | 74,500 | 49.00 | 38,000 |
| SILGA | 25,000 | 61,000 | 40.98 | 36,000 |
| Yenagoa | 56,000 | 91,254 | 62.23 | 35,254 |

**Figure 3.1:**
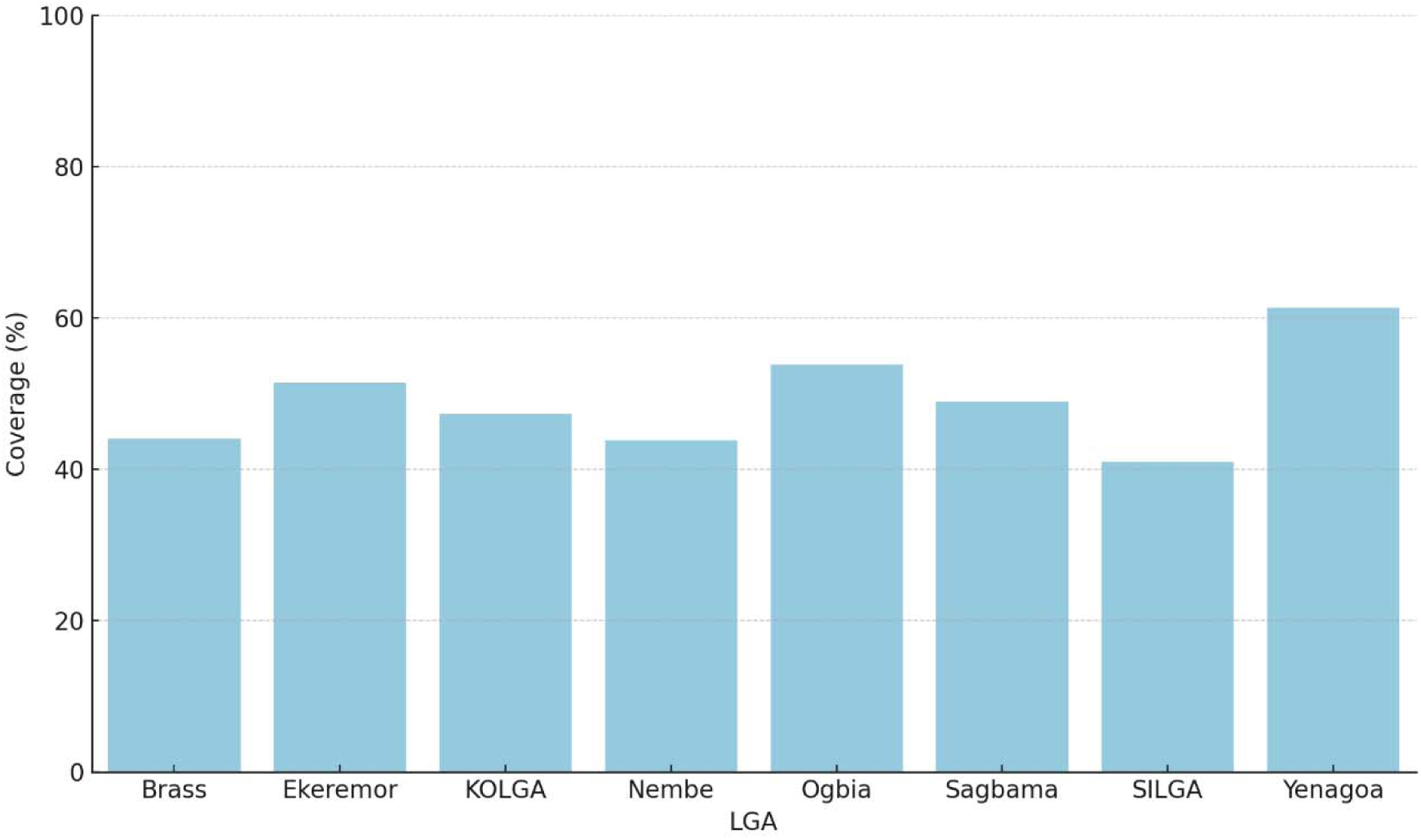
Bar Chart Showing Deworming Coverage (%) by LGA.

**Figure 3.2:**
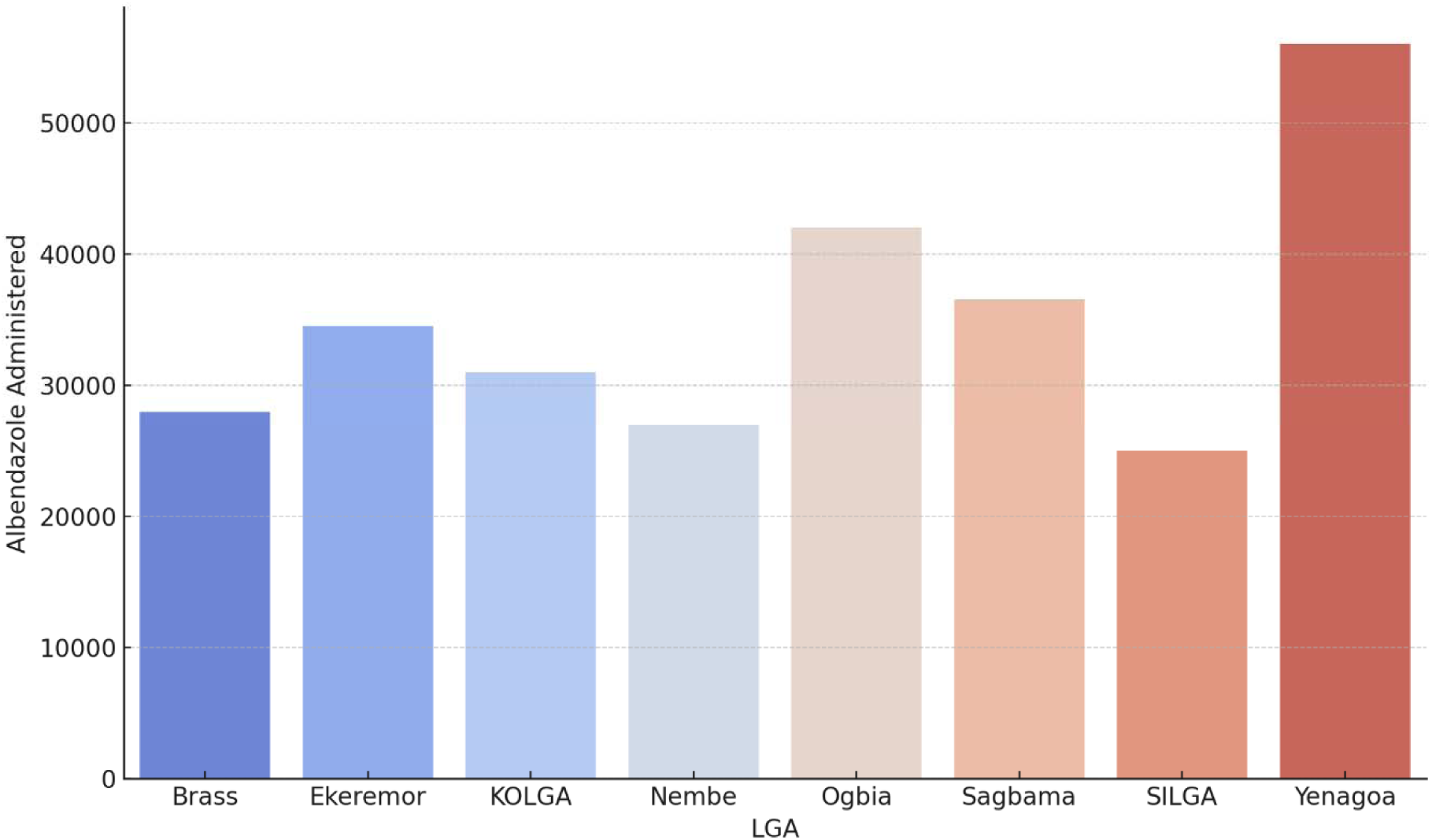
Map of Deworming Reach per LGA Using Heat Zones.

### 3.2 Comparison Between Deworming and Vitamin A Supplementation

To assess the balance of program delivery, deworming coverage was compared with that of vitamin A supplementation another key intervention delivered during the MNCH Week. The aim was to identify service delivery gaps and prioritize interventions for future campaigns.

As illustrated in Table 3.2, every LGA recorded higher coverage for vitamin A supplementation compared to deworming. For example, in Ogbia, 42,000 children were dewormed compared to 61,000 who received vitamin A, showing a coverage difference of 24.36 percentage points. Yenagoa had the highest vitamin A reach (82.86%), while also leading in deworming, though with a lower gap (20.63 percentage points). SILGA again had the weakest performance on both indicators, but the vitamin A reach (57.38%) still exceeded its deworming coverage by over 16 points.

**Table 3.2:** Comparison of Deworming and Vitamin A Coverage by LGA.

| LGA | Deworming | Vitamin A | Coverage Gap | Vitamin A Not |
| --- | --- | --- | --- | --- |

|  | Coverage (%) | Coverage (%) | (%) | Received |
| --- | --- | --- | --- | --- |
| Brass | 44.09 | 63.78 | 19.69 | 23,000 |
| Ekeremor | 51.49 | 71.64 | 20.15 | 19,000 |
| KOLGA | 47.33 | 67.18 | 19.85 | 21,500 |
| Nembe | 43.90 | 61.79 | 17.89 | 23,500 |
| Ogbia | 53.85 | 78.21 | 24.36 | 17,000 |
| Sagbama | 49.00 | 65.77 | 16.77 | 25,500 |
| SILGA | 40.98 | 57.38 | 16.40 | 26,000 |
| Yenagoa | 62.23 | 82.86 | 20.63 | 15,647 |

**Figure 3.3:**
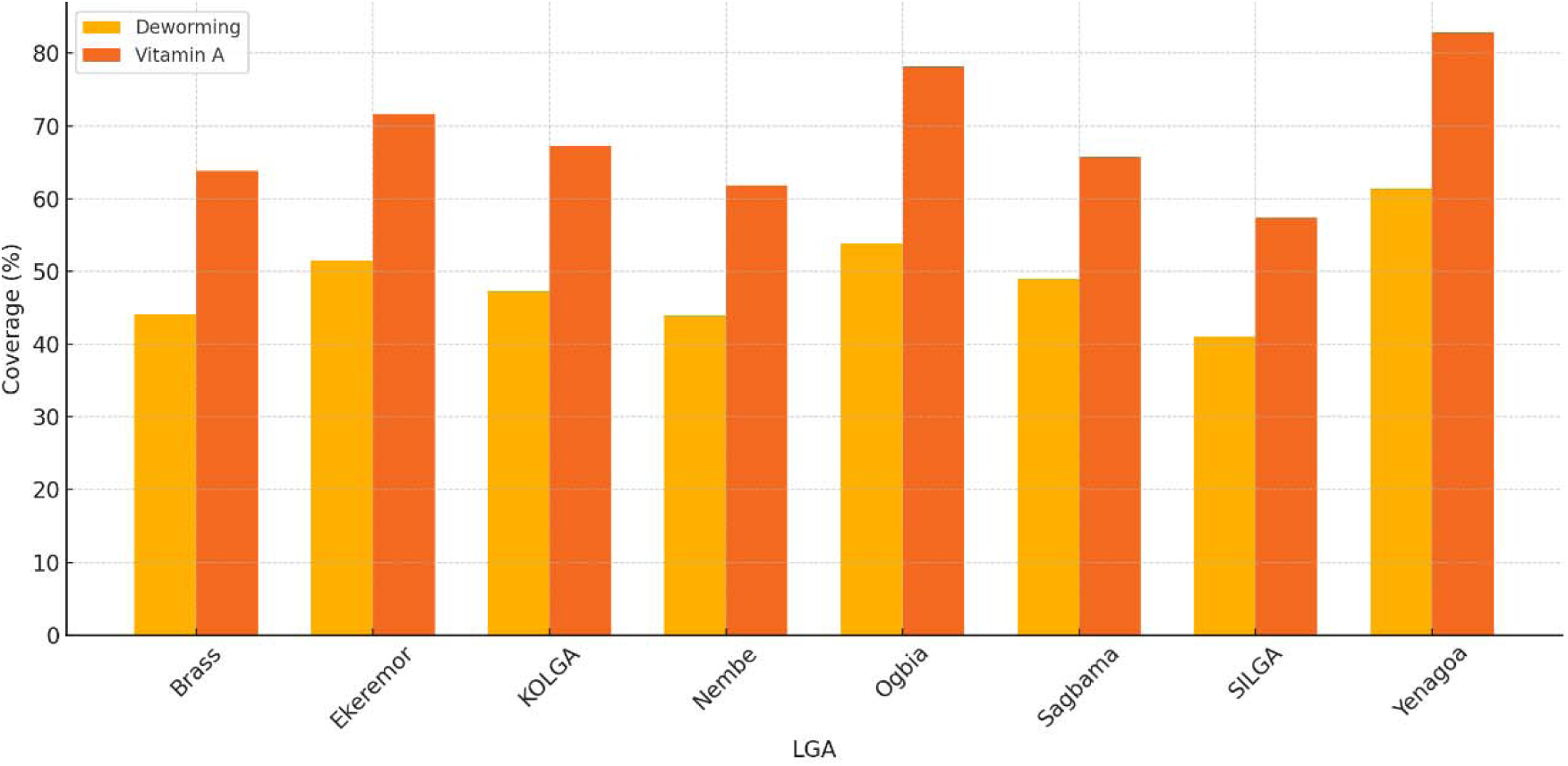
Side-by-Side Bar Chart Comparing Deworming and Vitamin A Coverage.

**Figure 3.4:**
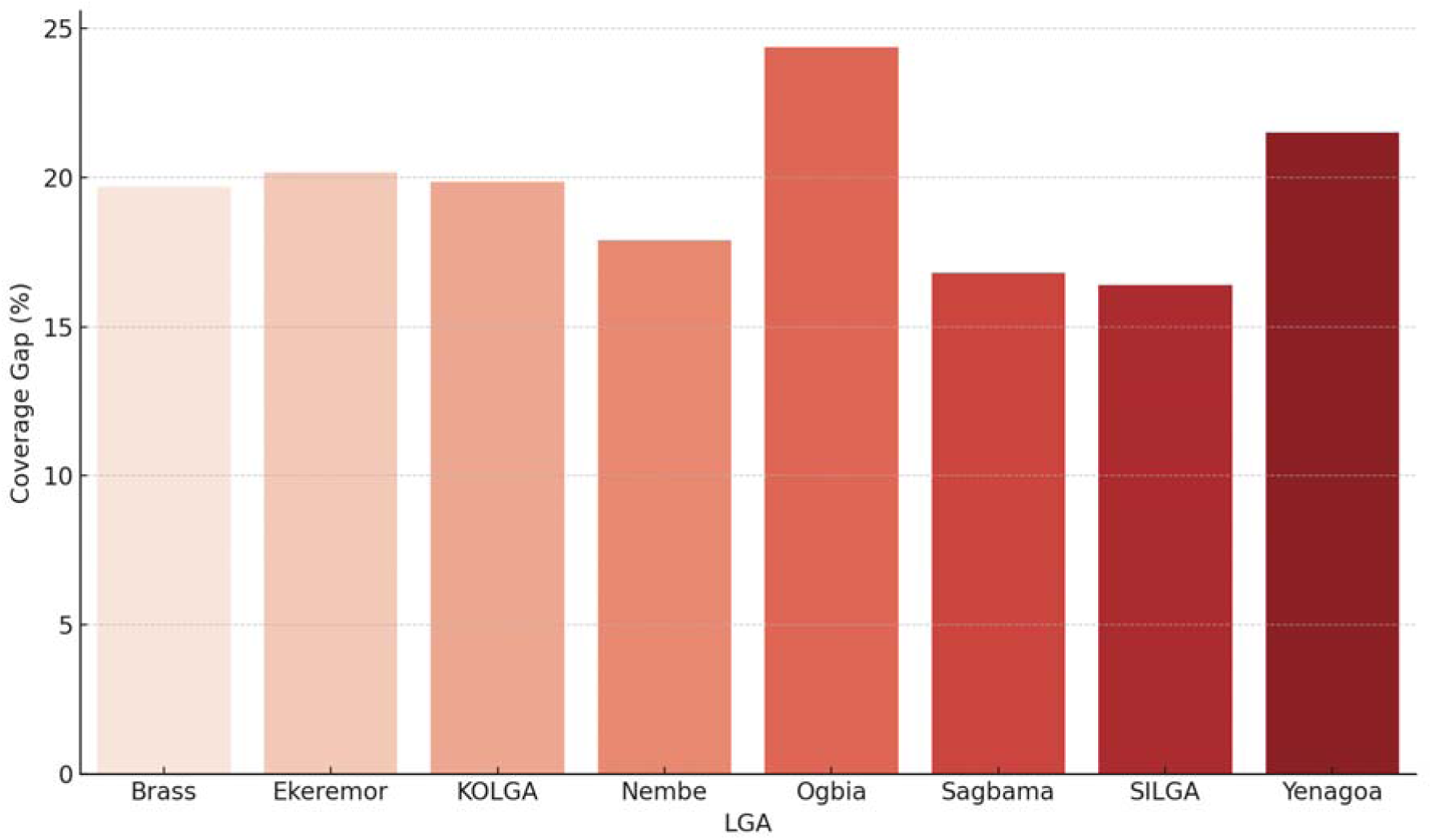
Gap Chart Showing Difference Between Deworming and Vitamin A per LGA.

### 3.3 State-Level Summary of Deworming and Vitamin A Outcomes

Table 3.3 aggregates the coverage statistics for all LGAs to present an overall picture of MNCH Week performance across Bayelsa State. Out of a projected 562,254 eligible children, 280,000 received deworming and 391,107 received vitamin A. This indicates that approximately 49% of the target population received Albendazole, while nearly 69% received vitamin A.

The mean deworming coverage across LGAs was 49.0%, significantly lower than the mean vitamin A coverage of 68.6%. The lowest deworming coverage was recorded in SILGA (40.98%) and the highest in Yenagoa (62.23%).

**Table 3.3:** Statewide Deworming and Vitamin A Summary Statistics.

| Metric | Value |
| --- | --- |
| Total Target Children | 562,254 |
| Total Dewormed | 280,000 |
| Total Given Vitamin A | 391,107 |
| Mean Deworming Coverage (%) | 49.00 |
| Mean Vitamin A Coverage (%) | 68.57 |
| Lowest Deworming Coverage (%) | 40.98 |
| Highest Deworming Coverage (%) | 62.23 |

**Figure 3.5:**
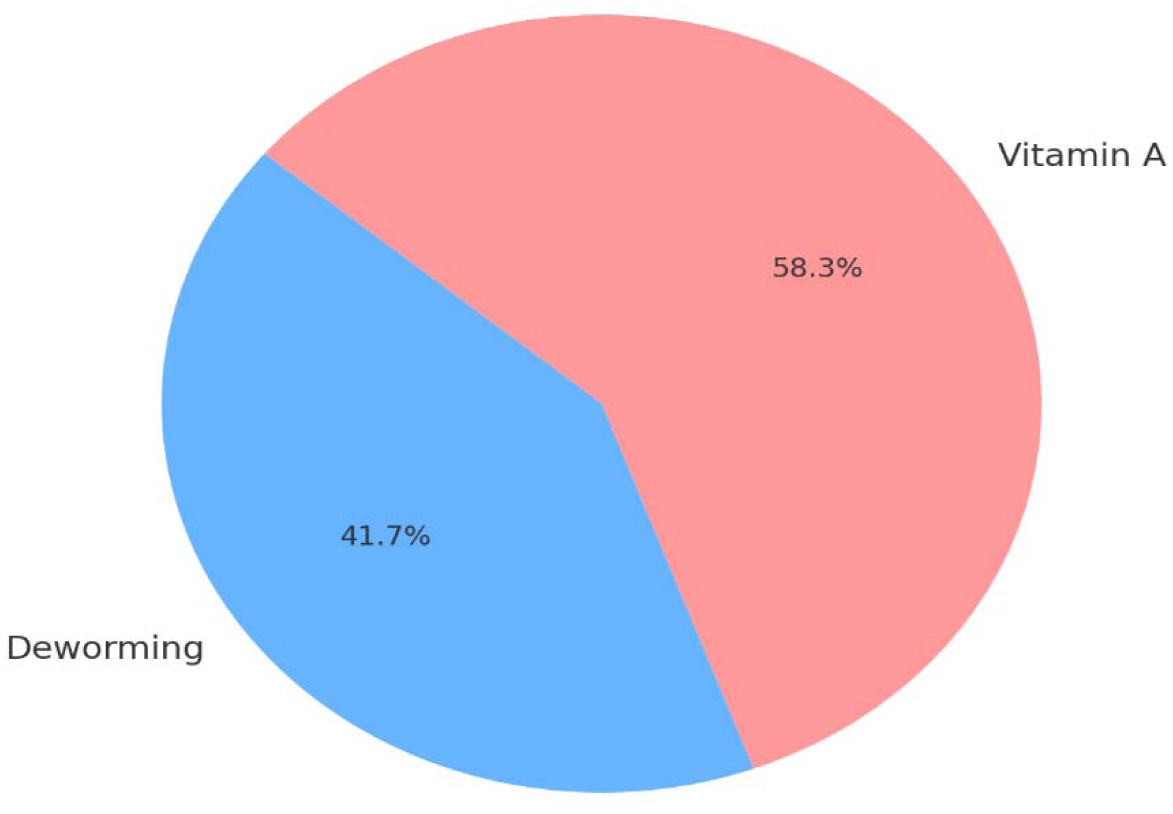
Pie Chart Showing State-Level Distribution of Vitamin A and Deworming Coverage.

**Figure 3.6:**
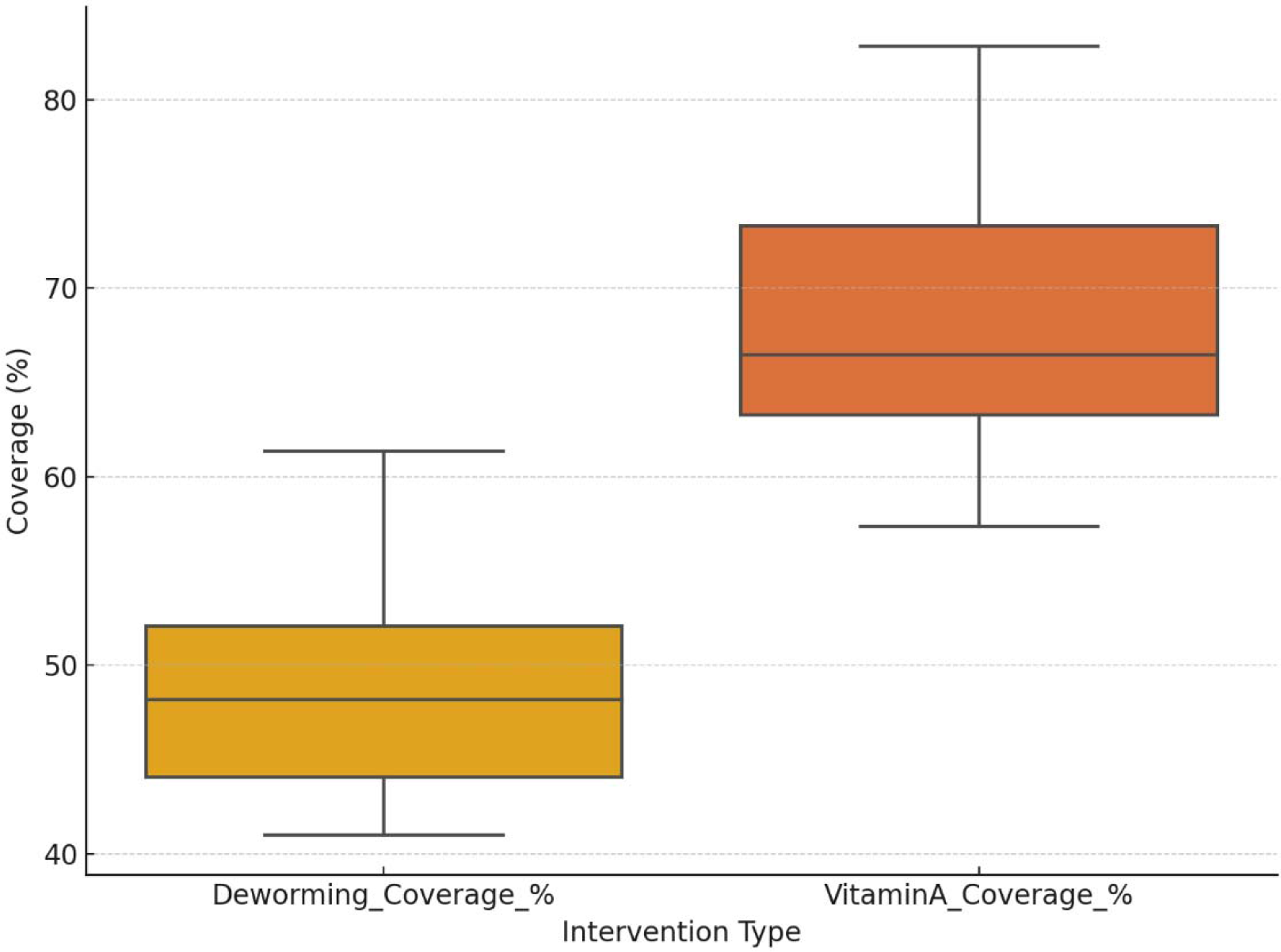
Boxplot Comparing LGA Coverage Distribution for Both Interventions.

## CHAPTER FOUR: DISCUSSION OF FINDINGS

### 4.1 Deworming Coverage Across LGAs

The analysis of Albendazole administration across the eight LGAs of Bayelsa State presents a clear picture of systemic disparities in service reach. Despite a unified campaign strategy, the actual reach of deworming interventions was highly uneven, with coverage percentages ranging from as low as 40.98% in Southern Ijaw (SILGA) to 62.23% in Yenagoa. These figures are particularly significant because they illustrate how state-wide planning often struggles to translate into equitable coverage across different regions, especially those with challenging terrain or weaker health infrastructure.

The relatively higher coverage in Yenagoa is not surprising. As the state capital and most urbanized LGA, it benefits from denser health facility coverage, better transport networks, and easier community access. Health workers stationed here are also more likely to receive training promptly, have access to supplies on time, and benefit from efficient supervision. These factors contribute directly to better campaign outcomes. In contrast, LGAs like Brass, SILGA, and Nembe have difficult terrain, riverine settlements, and lower population densities, all of which complicate last-mile delivery and community mobilization. These areas often face delays in supply delivery, staffing shortages, and communication gaps, which limit their ability to meet deworming targets.

Another point of concern is the sheer volume of children who did not receive deworming medication. In nearly every LGA, the number of uncovered children exceeded 30,000, representing a significant missed opportunity in terms of both immediate parasite control and long-term health outcomes. These gaps point to potential inefficiencies in enumeration, resource allocation, and ward-level implementation. The reliance on manual planning tools, potential inconsistencies in population estimates, and limited microplanning support for hard-to-reach communities may have further contributed to this outcome.

Additionally, stakeholder and staff readiness plays a role. While deworming was part of the integrated package, it may not have been emphasized as strongly as other interventions. If frontline workers were insufficiently trained or lacked confidence in the proper administration of Albendazole, it could result in cautious under-delivery or inaccurate reporting. Moreover, the community awareness component might not have been robust enough to educate caregivers about the importance of deworming, leading to reluctance or apathy in attendance.

The data make it evident that coverage challenges are not merely logistical but also systemic. LGAs with better historical performance, access to experienced staff, and smoother logistics consistently performed better, revealing structural inequities in service delivery. Unless these issues are addressed through LGA-specific interventions, infrastructure support, and stronger accountability mechanisms, coverage outcomes will continue to reflect the state’s uneven development.

### 4.2 Comparison Between Deworming and Vitamin A Supplementation

A compelling insight from the campaign performance data is the persistent gap between deworming and vitamin A coverage. Across all LGAs, vitamin A outperformed Albendazole in coverage sometimes by as much as 24 percentage points. This consistent trend demands closer examination of program execution strategies, community engagement, and health worker behavior.

The question arises: why does vitamin A enjoy better coverage than deworming, even when both are delivered under the same conditions, by the same workers, to the same children? One explanation could lie in caregiver perception. Vitamin A has been administered in Nigeria for decades and is often associated with visual health and immunity concepts that caregivers may be more familiar with. In contrast, deworming, though critically important, may not be well understood by parents, especially in rural or lower-literacy communities. The term "deworming" might not carry the same urgency or emotional appeal as "vitamin A to protect your child’s eyes or prevent disease."

There may also be operational factors. Vitamin A tablets are typically smaller, easier to handle, and less likely to require follow-up or observation post-administration. Albendazole, on the other hand, must be administered with certain caution, particularly in cases where children may have eaten little or may be at risk of side effects. Health workers may have defaulted to prioritizing the “easier” intervention, especially under pressure or in time-constrained outreach settings. This type of triaging, though unintentional, results in lopsided service delivery.

The figures also point to a planning imbalance. If vitamin A tablets were distributed to health facilities or STF teams more reliably than Albendazole perhaps due to supply delays, miscommunication, or poor storage conditions then the coverage difference would emerge not from human behavior but from commodity availability. In large-scale campaigns, a delay of even a single day in receiving deworming supplies could result in several thousand missed children, particularly in wards with short windows of operation.

Moreover, there is the possibility that recording practices differ. If vitamin A was recorded with greater accuracy or with stronger emphasis in the service delivery forms, it might reflect better in the final report compared to deworming. Some STF teams may have failed to complete all columns in the deworming registers or may have merged it with other indicators, resulting in underreporting. This raises questions about the data collection protocols and how closely they were monitored at LGA and state levels.

Ultimately, the consistent superiority of vitamin A coverage across all LGAs reflects an institutional and social bias in health programming that needs to be addressed. Campaign equity is not just about whether services are offered, but whether they are delivered with equal strength, visibility, and support. Future MNCH Weeks must pay closer attention to the mechanisms behind service imbalance whether logistical, behavioral, or perceptual to ensure that all components of the child health package receive equal commitment.

### 4.3 State-Level Performance and Program Imbalance

At the macro level, Bayelsa State’s campaign data presents a mixed report card. With 562,254 children targeted during the MNCH Week, only about 49% received Albendazole, compared to 69% who received vitamin A supplementation. This gap, though already significant at the LGA level, becomes even more pronounced when aggregated across the state. It suggests that the deworming intervention, despite its inclusion in planning documents and training manuals, may not have received the operational support and on-the-ground prioritization needed for optimal execution.

The statewide mean coverage difference nearly 20 percentage points exposes a strategic imbalance in implementation. While it is commendable that nearly 400,000 children received vitamin A, the fact that over 280,000 children missed deworming raises serious concerns. These are children who remain exposed to helminth infections that impair nutrition, growth, and cognitive development. At a population level, the missed coverage translates to a delayed or diminished return on investment for broader public health goals.

The boxplot analysis of coverage distributions further reveals inconsistencies. While the range of vitamin A coverage shows more fluctuation, it still consistently surpasses the deworming coverage in each LGA. This pattern reflects that even in lower-performing areas, vitamin A reaches more children than Albendazole. One interpretation is that vitamin A has stronger structural enablers perhaps through better supply chains, clearer roles for staff, or even stronger community routines. Another is that deworming continues to be treated as a secondary priority in health outreach, even when program guidelines emphasize its inclusion.

The pie chart of total intervention reach is perhaps the most visual representation of this imbalance: vitamin A occupies a larger share of the service distribution. This unevenness is not merely a numerical artifact it has real implications. If integrated campaigns continue to deliver some services well while underperforming in others, they risk creating partial health gains, weakening the overall impact of the intervention mix.

This pattern also suggests the need for better monitoring systems. Without real-time tracking of each service component, program managers may be unaware of underperformance until the campaign ends. By that point, missed opportunities cannot be recovered. Deploying mobile dashboards, structured feedback loops, and real-time alerts for commodity stock-outs or team inactivity can help rebalance performance across interventions.

Overall, Bayelsa’s MNCH Week data confirms that integration alone is not enough. Integrated delivery must be accompanied by integrated accountability, equitable training, and synchronized supply flows. The true success of such campaigns lies not just in how many children were reached, but in whether all children received all the interventions they were promised.

## CHAPTER FIVE: CONCLUSION AND RECOMMENDATIONS

### 5.1 Summary of Key Insights

The findings from this study have revealed more than just numerical gaps in health service delivery they have exposed the fundamental dynamics of how equity, execution, and emphasis shape outcomes in large-scale public health interventions. The June 2025 Maternal and Child Health Week in Bayelsa State was designed to be a transformative vehicle for child survival, bundling together essential services such as deworming, vitamin A supplementation, iron-folate distribution, MUAC screening, and birth registration. Yet, despite the integration of these life-saving services into a single outreach platform, the outcomes were uneven, particularly in the coverage of deworming.

With only 49% of the target population receiving Albendazole, in contrast to 69% who received vitamin A, the disparity is not only stark but also instructive. It underscores a critical lesson: integration in planning does not automatically ensure integration in execution. Deworming was evidently underprioritized in training, logistics, community sensitization, and perhaps most importantly, in the mindset of both implementers and beneficiaries.

### 5.2 Strategic and Operational Implications

The consistent underperformance of deworming, despite being paired with higher-performing interventions like vitamin A supplementation, signals significant operational bottlenecks. The fact that vitamin A was consistently delivered more widely under the same campaign conditions reveals gaps not in access but in coordination and emphasis. Structural and logistical factors ranging from commodity pre-positioning to LGA-level readiness played decisive roles in determining which interventions reached children and which did not.

Moreover, underperformance was not randomly distributed. LGAs with challenging geography and weaker infrastructure reported the lowest deworming coverage, reaffirming the correlation between systemic development gaps and health service inequity. Such results reflect the urgent need to move beyond one-size-fits-all planning and toward a differentiated model that accounts for contextual differences across locations.

### 5.3 The Case for Balanced Integration

Bayelsa’s MNCH Week has proven that with strong campaign structures, health systems can scale essential services rapidly. But it has also shown that without equal operational discipline across all intervention lines, campaigns can become functionally imbalanced. This threatens not only the effectiveness of individual services like deworming but also the trust and expectations of communities who believe they are receiving a comprehensive care package.

The results highlight the need to build integration not just into campaign logistics but into campaign values and accountability systems. The goal should not be merely to deliver several interventions together, but to deliver them with equal strength, visibility, and operational rigor. Deworming must no longer be treated as an add-on it must be embedded into the very backbone of child survival programming.

### 5.4 Recommendations for Strengthening Deworming Implementation

To chart a new course, a series of decisive steps must be taken bold, practical, and immediate:

- **Institutional Priority and Performance Tracking**: Deworming coverage should be tracked as a high-level KPI, on par with immunization and vitamin A. It must feature prominently in national dashboards, supervision checklists, and routine reviews.
- **Real-Time Monitoring Systems**: Mobile dashboards should be deployed during campaigns to track coverage by intervention, flagging real-time shortfalls in Albendazole delivery and enabling mid-week corrective action.
- **Commodity Chain Improvements**: Albendazole should have a distinct supply pipeline with clear delivery timelines, pre-positioning protocols, and accountability at every handoff.
- **Focused Staff Training**: Health workers must be retrained with an explicit focus on deworming its benefits, administration guidelines, and common misconceptions among caregivers.
- **Tailored Microplanning Tools**: Each intervention should have its own microplanning template to prevent unintentional under-documentation or neglect during field execution.
- **Community-Level Advocacy**: Engage faith leaders, town announcers, women’s groups, and youth networks to demystify deworming and generate demand that matches that of vitamin A.
- **Support for Low-Performing LGAs**: Introduce a tiered support system where LGAs with chronic underperformance receive additional staffing, extended campaign periods, and increased mobilization funding.

### 5.5 Concluding Reflections

In summary, this study is not merely a performance report it is a call to action. It reminds us that the value of integration lies not in what is planned, but in what is equally delivered. The health of a child in Nembe should not be compromised because of the terrain, just as the importance of Albendazole should not be overshadowed by the popularity of vitamin A. The gaps identified are not insurmountable but they demand intentional correction.

Bayelsa State and by extension, Nigeria now stands at a point of opportunity. The tools exist, the commitment is present, and the need is undeniable. To protect every child, to fulfill the promise of health equity, and to uphold the credibility of integrated campaigns, it is time to act with urgency and precision. Let every future MNCH Week be not only broader in reach but deeper in impact where every child, everywhere, receives every intervention they deserve.

## Data Availability

All data produced in the present work are contained in the manuscript

